# SARS-CoV-2 infection risk during delivery of childhood vaccination campaigns: a modelling study

**DOI:** 10.1101/2021.05.14.21257215

**Authors:** Simon R Procter, Kaja Abbas, Stefan Flasche, Ulla Griffiths, Brittany Hagedorn, Kathleen M O’Reilly, CMMID COVID-19 Working Group, Mark Jit

## Abstract

**Background:** The COVID-19 pandemic has disrupted delivery of immunisation services globally. Many countries have postponed vaccination campaigns out of concern about infection risks to staff delivering vaccination, the children being vaccinated and their families. The World Health Organization recommends considering both the benefit of preventive campaigns and the risk of SARS-CoV-2 transmission when making decisions about campaigns during COVID-19 outbreaks, but there has been little quantification of the risks.

**Methods:** We modelled excess SARS-CoV-2 infection risk to vaccinators, vaccinees and their caregivers resulting from vaccination campaigns delivered during a COVID-19 epidemic. Our model used population age-structure and contact patterns from three exemplar countries (Burkina Faso, Ethiopia, and Brazil). It combined an existing compartmental transmission model of an underlying COVID-19 epidemic with a Reed-Frost model of SARS-CoV-2 infection risk to vaccinators and vaccinees. We explored how excess risk depends on key parameters governing SARS-CoV-2 transmissibility, and aspects of campaign delivery such as campaign duration, number of vaccinations, and effectiveness of personal protective equipment (PPE) and symptomatic screening.

**Results:** Infection risks differ considerably depending on the circumstances in which vaccination campaigns are conducted. A campaign conducted at the peak of a SARS-CoV-2 epidemic with high prevalence and without special infection mitigation measures could increase absolute infection risk by 32% to 45% for vaccinators, and 0.3% to 0.5% for vaccinees and caregivers. However, these risks could be reduced to 3.6% to 5.3% and 0.1% to 0.2% respectively by use of PPE that reduces transmission by 90% (as might be achieved with N95 respirators or high-quality surgical masks) and symptomatic screening.

**Conclusions:** SARS-CoV-2 infection risks to vaccinators, vaccinees and caregivers during vaccination campaigns can be greatly reduced by adequate PPE, symptomatic screening, and appropriate campaign timing. Our results support the use of adequate risk mitigation measures for vaccination campaigns held during SARS-CoV-2 epidemics, rather than cancelling them entirely.

## Background

The Coronavirus Disease 2019 (COVID-19) pandemic, caused by the severe acute respiratory syndrome coronavirus 2 (SARS-CoV-2), has severely disrupted healthcare service delivery globally. In a pulse survey of key informants between May and July 2020, respondents from 90% of the surveyed countries reported disruptions to essential health services, with the greatest disruptions reported in low- and middle-income countries (LMICs)^1^. The same survey reported that one of the most severely disrupted services has been delivery of both outreach and facility-based immunisation.

This disruption has resulted from multiple factors including interruption of supply chains, limitations on travel, and diversion of finances and healthcare workers due to COVID-19 and the associated response. One important reason for the disruption has been concern around the potential for SARS-CoV-2 transmission during provision of immunisation, particularly through delivery of vaccination campaigns^2^. There is particular concern over putting vaccination staff at increased risk of COVID-19, since healthcare workers are already at high risk of COVID-19^3^, and healthcare workforce pressures are particularly acute due to the need to care for COVID-19 patients.

In March 2020, at the start of the COVID-19 pandemic, the World Health Organization (WHO) and Pan American Health Organisation (PAHO) recommended temporary suspension of preventative immunisation campaigns but encouraged continuation of routine immunisation^2,4^. In November 2020, with the first COVID-19 wave abating in many LMICs, WHO issued more nuanced recommendations encouraging countries to evaluate decisions around vaccine campaigns by considering both the risk of disease from missed vaccine doses, and the risk of SARS-CoV-2 transmission during campaigns^5^. Some countries (e.g. Ethiopia, DRC and Somalia) that had previously cancelled and postponed vaccine campaigns had reinstated them by September 2020^6–8^. Cancellation of vaccination campaigns have also impacted global efforts to eradicate polio, which is itself an ongoing public health emergency of international concern^9^.

However, quantitative evidence about SARS-CoV-2 infection risk during vaccination campaigns is limited, and urgently needed as many countries face new COVID-19 waves in 2021. Previous modelling studies have demonstrated that the benefits of continuing routine immunisation likely outweigh the excess risk from COVID-19 but did not examine campaign delivery^10^. A recent study has examined risks of vaccine-preventable disease outbreaks (measles, meningococcal A, and Yellow Fever) associated with delaying immunisation campaigns, which varied across countries^11^. Another study assessed the risk of measles outbreaks in Kenya and found that although COVID-19 interventions also temporarily reduced the risk of an outbreak from measles immunity gaps, this risk rises rapidly once these restrictions are lifted highlighting the need to implement catch-up campaigns^12^. One study looked at the risk of transmission in the community during fixed-post or house-to-house immunisation campaigns in six countries (Angola, Ecuador, Pakistan, Ukraine, Nepal and Lao PDR), but did not model specific interactions between vaccination staff and service users^13^.

To address this evidence gap, this study models the additional risk of SARS-CoV-2 infection to children receiving vaccination (hereinafter vaccinees), their accompanying caregivers, and vaccinators delivering either fixed-post or house-to-house vaccination campaigns during a simulated COVID-19 epidemic. Our analysis uses demographics from three exemplar countries (Burkina Faso, Ethiopia, and Brazil), and explores which factors are most important in determining the magnitude of these infection risks.

## Methods

We developed a modelling framework that combines an existing compartmental transmission model of SARS-CoV-2 (CovidM) with a novel mathematical model of SARS-CoV-2 infection risk during campaigns^14^. The transmission model was used to simulate different epidemic scenarios in the general population at a national level (i.e. without regional stratification) in different country settings. We then examined the additional infection risk to vaccinators, vaccinees and their caregivers, of immunisation campaigns conducted at different points during the epidemic. We also explored how this risk depends on key aspects of campaign delivery such as duration of the campaign, number of children vaccinated, and effectiveness of personal protective equipment (PPE) in reducing transmission.

### SARS-CoV-2 transmission model

We performed deterministic simulations of the SARS-CoV-2 epidemic using CovidM, which uses an age-stratified compartmental SEIR structure (Susceptible, Exposed, Infected - with sub-compartments for asymptomatic, pre-symptomatic and symptomatic infection - and Recovered) and has been described in detail by Davies et al.^14^ We use the same values as Davies et al. for epidemiological parameters including the latent period, and duration of sub-clinical, pre-clinical, and clinical infectiousness. We also make the same assumptions about the probability of developing clinical symptoms among different age groups^15^. Our simulations assumed no waning of natural immunity.

To explore the potential impact of population age-structure and social contact patterns on our results we parameterised our model using data for three exemplar countries: Burkina Faso, Ethiopia, and Brazil (Additional file 1: Fig. S1 and Fig. S2). We used population data from the United Nations population estimates^16^, and for social contact patterns utilised country- and age-specific synthetic contact matrices reported by Prem et al.^17^ These countries were chosen based on having respectively the lowest, median, and highest population median age amongst countries with a measles vaccination campaign planned for 2020^18^ (but excluding high-income countries and countries for which social contact matrices were unavailable). Our objective was not to predict the actual SARS-CoV-2 epidemics experienced by these particular countries, but rather to generate plausible scenarios using alternative demography and contact patterns to examine the implications for the risk associated with vaccination campaigns.

For our base case analysis, we modelled an epidemic with an R_0_ of 2, and explored alternative values as a sensitivity analysis (Table 1). A meta-analysis of the literature reported a mean R_0_ of 2.6 (SD 0.5)^19^; however we chose a lower value to reflect the impact of measures such as hand hygiene, mask-wearing and improved indoor ventilation on community transmission. We additionally assumed a base case reduction of 40% in all non-household contacts (i.e. school, work, and other settings) to reflect the impact of physical distancing policies, and varied these parameters in sensitivity analyses. These assumptions were chosen to generate partially mitigated epidemics that persist over a period of many months. Such scenarios are likely to be most challenging from a decision-making perspective. If mitigation is much stronger the epidemic will be completely suppressed, and conversely with little mitigation most of the population will become infected; in either case vaccination campaigns are likely to represent little additional risk.

**Table 1.** Model parameters used in base case and one-way sensitivity analyses.

| Parameter | Description | Base case | Sensitivity Analysis | Source |
| --- | --- | --- | --- | --- |
| $R_0$ | Basic reproduction number | 2.0 | 1.8, 2.2 | Assumed |
|  | Age-specific contact matrices | See Additional file 1: Fig S2. |  | 17 |
|  | Reduction in contacts outside of home due to NPIs | 40% | 20%, 60% (30%, 20%)* | Assumed |
| $u$ | Susceptibility to infection on contact | Derived from $R_0$ as described in Davies et al. | | 14 |
| $d^e$ | Latent period (pre-infectious) | 4 days | 2 days, 6 days | 147/21/21<br>12:40:00 PM |
| $d^r$ | Duration of infectiousness | 5 days | 3 days, 7 days | 14 |
| $d^c$ | Duration of vaccination campaign | 10 days | 5 days, 20 days^ | Expert opinion + 8,23 |
| $v$ | Number of children vaccinated by a vaccinator per day | Fixed-post: 150<br>House-to-house: 75 | Fixed-post: 75, 300<br>House-to-house: not varied | Expert opinion + 8,23,247/21/2<br>1 12:40:00 PM |
| $n^{cj}$ | Number of community contacts of children / caregiver during trip to / from vaccine clinic | Fixed-post: 5<br>House-to-house: 0 | Fixed-post: 1, 10<br>House-to-house: not varied | Assumed |
| $n^{cv}$ | Number of vaccination staff contacted by children / caregivers during vaccination | 1 | 3 | Expert opinion + 267/21/21<br>12:40:00 PM |
| $n^{vj}$ | Number of extra community contacts of vaccinators for each household visited during campaign | Fixed-post: 0<br>House-to-house: 1 | House-to-house: 0, 2 | Assumed |
| $\pi$ | Effectiveness of PPE in reducing transmission | 75% | 0%, 50%, 90%, 100% | 22,27 |
| $\rho$ | Relative effectiveness of PPE in reducing transmission to vaccinators or vaccinees/caregivers | 1 | 0.5 for vaccinators, 0.5 for caregivers | Assumed |

\*This was used to model the assumption of faster epidemics amongst healthcare workers.
^We performed two separate sensitivity analysis for campaign duration: (i) $v$ was kept constant so the total number of vaccinations per vaccinator changed with campaign duration; (ii) $v$ was simultaneously adjusted so the total number vaccinations per vaccinator remained constant.

The model was run for a period of two years, with the epidemics seeded by a fixed number of individuals aged between 20 and 50 each day during the first week of the simulations; this number being chosen for each country such that the total number of infections seeded was 1 per 100,000 of the general population. The outputs from the simulations were then used to calculate the overall proportion of the population (across all age groups) on a given day *t* that is either susceptible (*s*_*t*_), exposed (*e*_*t*_), infected but asymptomatic 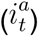, infected but pre-symptomatic 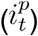, infected and symptomatic 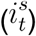, or recovered (*r*_*t*_). These outputs were then used as inputs into the vaccination campaign risk model (described below.)

### Vaccination campaign risk model

#### Risk to vaccinators

We first estimate the infection risk to vaccinators by extending our previous work^10^ on the risk associated with routine immunisation based on a Reed-Frost type model^20,21^. For a susceptible vaccinator, the individual risk of becoming infected by day *d* of a vaccination campaign that begins on day *t* of the epidemic is given by,

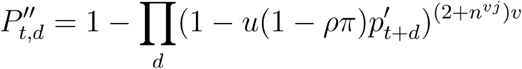

and the excess risk amongst all vaccinators accounting for those who are susceptible at the time of the campaign is,

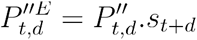

where the prevalence of infectious individuals amongst community members attending vaccination services is 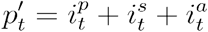 in scenarios without symptomatic screening, and 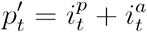 in scenarios with symptomatic screening; *u* is the transmissibility (probability of transmission per contact) of SARS-CoV-2; and *v* is the daily number of individuals vaccinated by the vaccinator. For symptomatic screening this assumes the maximum potential impact whereby the contribution of all symptomatic individuals to transmission is reduced to zero. For house-to-house campaigns vaccinators are assumed to contact an additional *n*^*vj*^ community members when travelling between households. The transmissibility *u* is governed by R_0_ and country-specific contact patterns, and was derived using CovidM by calculating the ratio of R_0_ to the dominant eigenvalue of the Next Generation Matrix (see supplementary appendix of Davies et al.^14^)

In the base case the fraction of vaccinators susceptible on day *t* in the absence of the campaign is assumed to be the same as for the general population,*s*_*t*_. We assume that the combined effect of PPE available to the vaccinators and other hygiene measures leads to a reduction by a factor π in transmission. In scenario analysis we explore the impact if the effect of PPE is lower (by a factor ρ) at protecting either vaccinators or vaccinees and caregivers, which might occur, for example, due to differences in type and/or use of PPE, or to different impacts on source control vs personal protection^22^.

#### Risk to vaccinees and their caregivers

Among susceptible individuals, the risk of either the vaccinee and/or their caregiver being infected on day of a campaign that begins on day of the epidemic is given by,

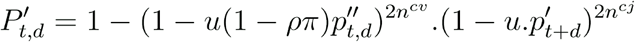

and the excess risk accounting for those who are susceptible is,

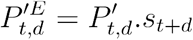

where *n*^*cj*^ is the number of community contacts during the visit to the vaccine clinic (assumed to be zero for house-to-house campaigns), 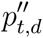 is the prevalence amongst vaccinators on day *d* of a campaign that began on day *t*, and all other terms have the same meaning as above. We account for the fact that vaccinators who are infected as a result of the campaign will not become infectious until after a period *d*^*e*^, and will subsequently recover after a period *d*^*r*^, by calculating the prevalence as follows,

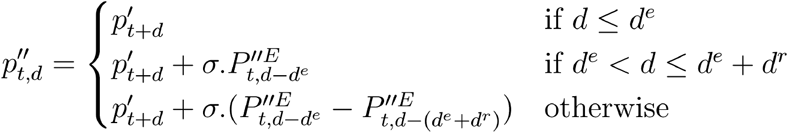

where the factor σ is set to σ =1 for scenarios without symptomatic screening, and for scenarios with symptomatic screening we assume that on a given day *d* the proportion of vaccinators infected as a result of the campaign who are symptomatic is the same as the symptomatic fraction in the general community such that 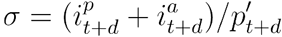.

### Model parameterisation

Model input parameters for the vaccination risk model are listed in Table 1. In our base case we assume that, prior to the vaccination campaign, vaccinators have the same baseline infection risk as the general population. However, we also investigated scenarios in which healthcare workers were assumed to experience a faster epidemic, and therefore have initially higher prevalence of infection, than the general population. This was modelled as an independent epidemic assuming a smaller reduction in out-of-home contacts, since healthcare workers must continue to work with the public. We assumed the same average duration for the latent and infectious periods as used in the transmission model.

To inform our assumptions about vaccine campaign characteristics we used a combination of literature values together with expert advice from a group of immunisation planning staff. According to WHO guidance, immunisation campaigns usually take place within a short timeframe lasting from a few days to about one month^23^. For our base case we modelled a campaign of 10 days duration, similar to a recent measles campaign in Ethiopia^8^, and varied this in sensitivity analyses from 5 to 20 days. We assumed that vaccinators would vaccinate an average of 150 children per day for fixed-post campaigns and 75 children per day in house-to-house campaigns^23,24^.

Evidence about social contacts relevant to infection transmission during healthcare visits is limited. One study in Singapore found that visitors to a hospital had a median of 2 contacts with healthcare workers (range 0-12) and 1 contact with other visitors (range 0-6)^25^. A time-and-motion study in India found that an immunisation clinic required caregivers and their children to come into contact with staff stationed at three separate tables within the clinic^26^. Therefore, we concluded that contacts during fixed-post immunisation visits are likely to be in single digits. For house-to-house campaigns, discussion with immunisation planning staff indicated that vaccinators are likely to have very few additional contacts, unless travelling in poor urban areas on foot. Our vaccination risk model implicitly assumes that the same meaning of a contact that underpins the contact matrices from Prem et al.^17^: either a physical contact or a two-way conversation in which 3 or more words were exchanged.

Hence for fixed-post campaigns, children and their caregivers were assumed to come into contact with an average of 5 additional people in the clinic waiting area, and during their journey to and from the vaccine clinic. In contrast, for house-to-house campaigns children and caregivers were assumed to have no additional community contacts, but vaccinators were assumed to have one additional community contact for each child vaccinated. To assess the impact of different parameter assumptions these were varied in sensitivity analysis.

For the effectiveness of PPE, cloth masks may reduce transmission by around 50-80%^22^, while N95 respirators can achieve even higher reductions^27^ but may not be available in all settings.

## Results

Our base case simulations resulted in epidemics that are similar across country settings (Fig. 1). In our base case scenario, with R_0_ = 2 and a 40% reduction in out-of-home contacts, the epidemic lasts for around one year with peak incidence of 0.23% to 0.26% per day, peak prevalence of 1.6% to 1.8%, and between 29% and 31% of the population infected by the end of the epidemic. We obtained very similar values when using demography and contact patterns for all three countries. Using alternative R_0_ values of 1.8 or 2.2 resulted in peak prevalence of 0.6% to 3.1%, and cumulative infections that ranged between 18% and 40%. A lower, 20%, reduction in contacts leads to higher peak prevalence (4.8 % to 5.1%), whereas 60% contact reduction results in suppression of the epidemic (Additional file 1: Fig. S3.)

**Figure 1.**
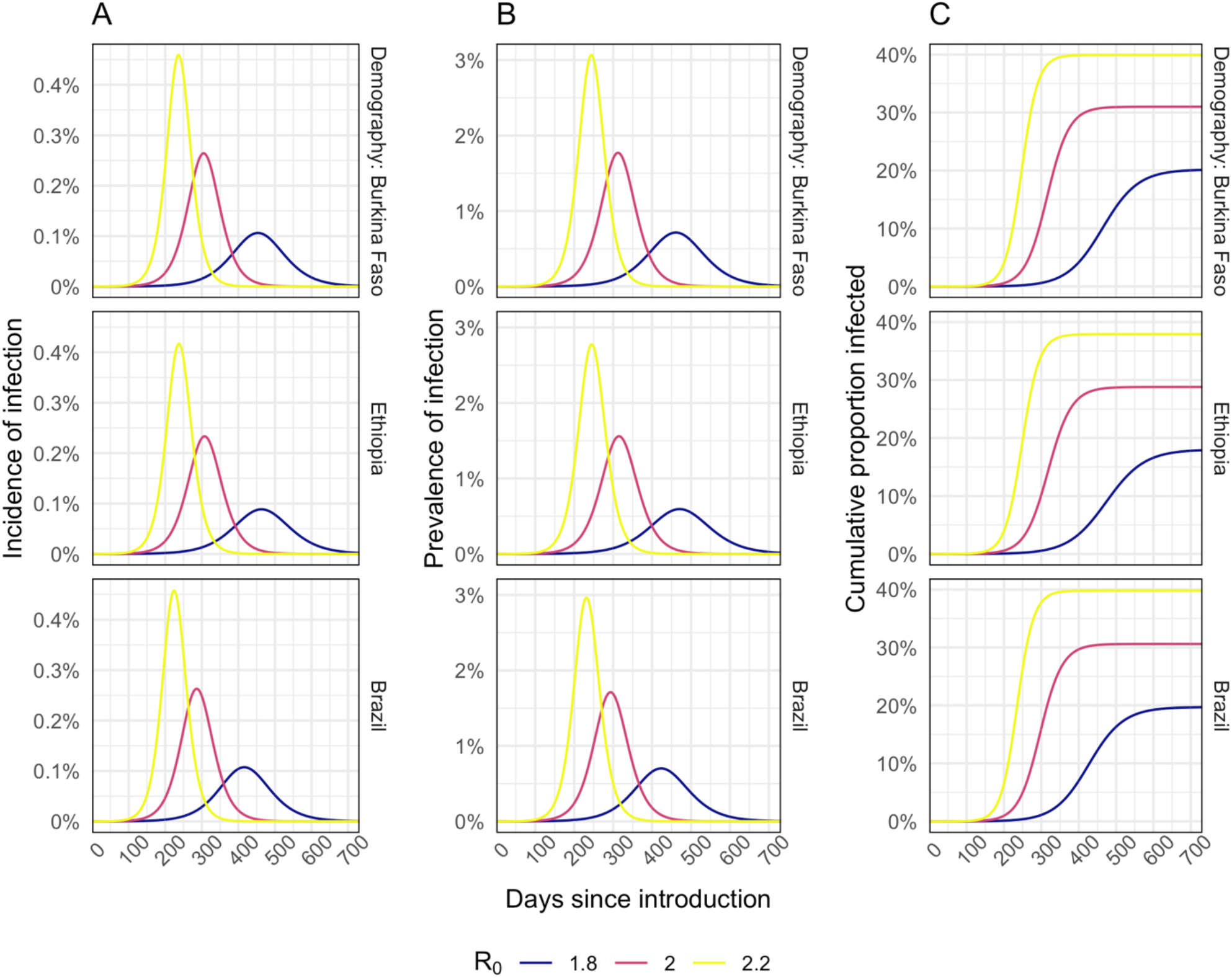
Modelled incidence (A), prevalence (B), and cumulative proportion of the population infected (C) for different R_0_ assumptions.

The prevalence of infection in the community (Fig. 1B and Additional file 1: Fig. S3) at the time of the campaign is a key driver of excess infection risk for both vaccinators (Fig. 2A and Additional file 1: Fig. S6A), and for children and their caregivers (Fig. 2B and Additional file 1: Fig. S6B). In the base case scenario peak excess risk of infection without effective PPE or symptomatic screening ranged from 32% to 45% for vaccinators and 0.30% to 0.54% for children and their caregivers. If PPE is 75% effective at preventing transmission these excess risks fall to 10% to 15%, and 0.15% to 0.25% respectively. Symptomatic screening alone, without effective PPE, only leads to a drop in excess infection risk of 2.3% to 4.5% for vaccinators and 0.03% to 0.08% for vaccinees and caregivers. With PPE that reduces transmission risk by 90% and symptomatic screening, vaccinators’ risks can be further reduced to 3.6% to 5.3%. However, for vaccine recipients the additional risk reduction is only slight (to 0.14% to 0.23%) since the remaining excess risk is dominated by other community contacts. Hence, providing even a moderate (50%) level of PPE for vaccinators may achieve most of the risk reduction for vaccinees and their caregivers (Fig. 2B.)

**Figure 2.**
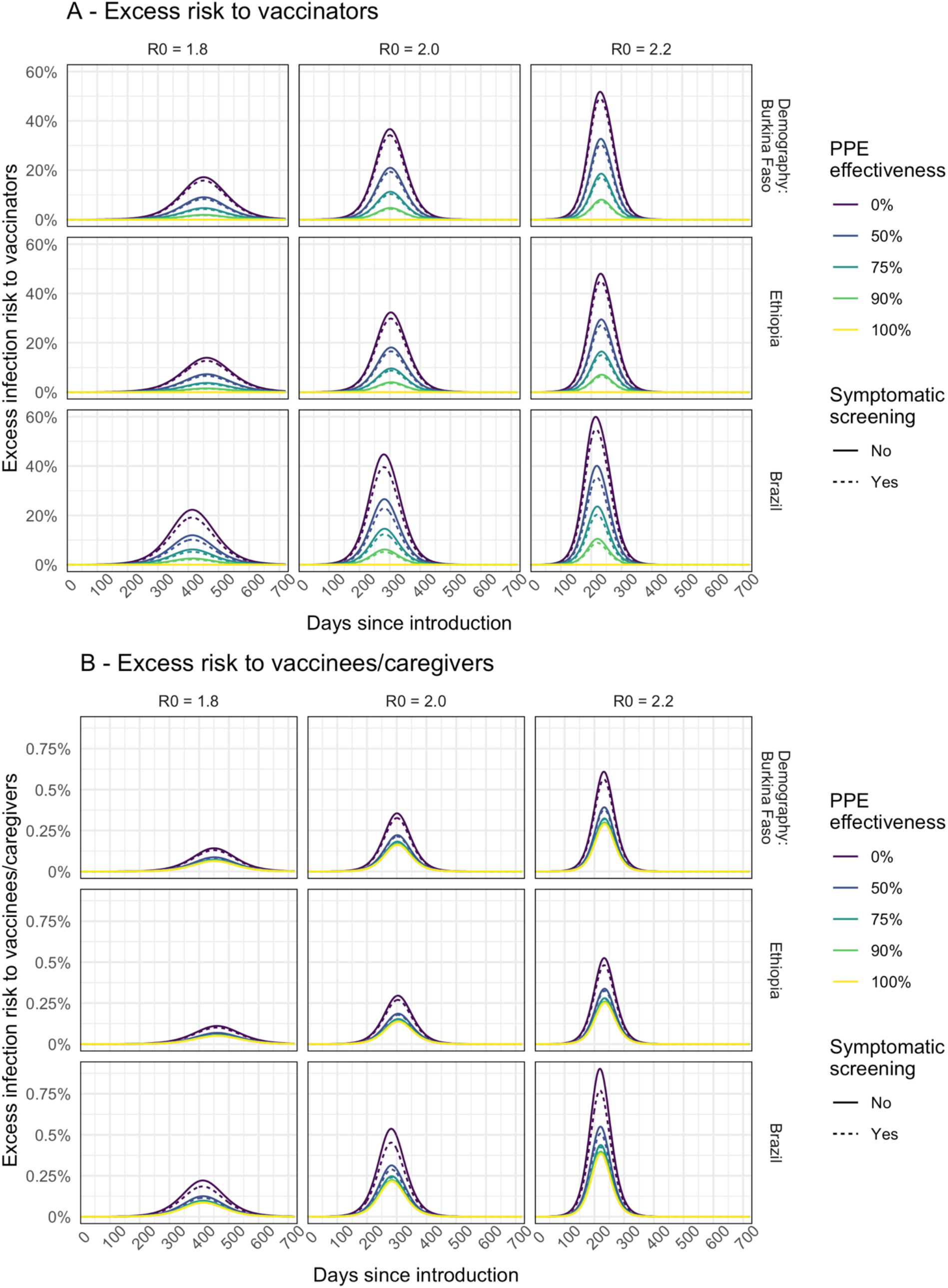
Modelled excess risk of (A) vaccinators and (B) children and/or caregivers becoming infected during fixed-post immunisation campaigns conducted at different times during the epidemic. Results are shown for epidemics modelled using different R_0_ assumptions. Line colour shows the impact of different levels of PPE effectiveness, and dashed lines show the combined impact of PPE together with symptomatic screening (assumed to screen out all symptomatic individuals.)

If healthcare workers are assumed to have a higher day-to-day risk of infection than the general population, then the baseline prevalence of infection among vaccinators (before the campaign) will be higher than that in the community during the earlier part of the epidemic (Additional file 1: Fig. S3). As a result, the excess infection risk for vaccinators due to the immunisation campaign is reduced (Additional file 1: Fig. S4A) because, compared to the base case, a lower proportion of vaccinators remain susceptible at a given point in the epidemic. For children and caregivers (Additional file 1: Fig. S4B) the impact is more nuanced. Excess infection risk is increased earlier in the epidemic since prevalence in vaccinators is higher and more children and caregivers are susceptible. But, later in the epidemic the excess risk can remain (relatively) high as prevalence rises among community contacts. However, this pattern is dependent on the extent of the lag between our modelled epidemics for the vaccinators and the general population.

For house-to-house delivery, compared to fixed-post delivery, under our base case assumptions, the excess risk is predicted to be lower both for the vaccinators (Additional file 1: Fig. S5A) and for vaccinees and caregivers (Additional file 1: Fig. S5B). For children the risk is lower compared to fixed-post delivery because there are assumed to be no additional community contacts that arise from travelling to or from a vaccine clinic. For the vaccinators the risk is lower because the number of children vaccinated per worker is lower, but if the number of people that vaccinators come into contact with during travel between households rises then the risk is increased.

In deterministic sensitivity analysis (Fig. 3) we found that for vaccinators, the peak excess infection risk was most sensitive to R_0_ (and the correspondingly higher peak in community prevalence) and to the total number of children vaccinated by each vaccinator during the campaign, which depends in turn on the number of vaccinations per day and the campaign duration. There is almost no impact of varying the campaign duration while holding the total number of vaccinations constant. Comparing scenarios with and without PPE, although this changes the overall excess risk, the model remains sensitive to the same parameters.

**Figure 3.**
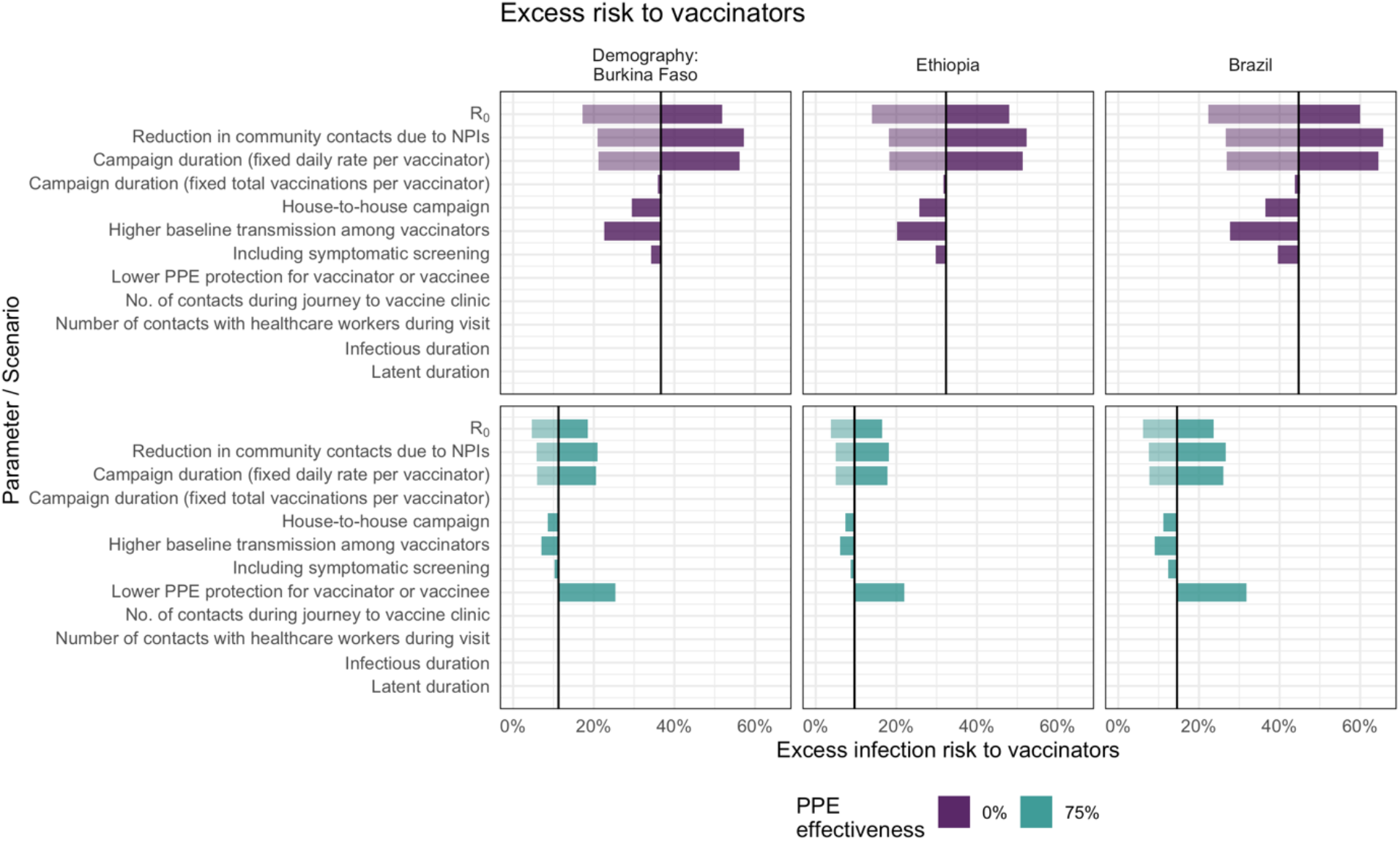
Sensitivity analysis showing the impact of varying model input parameters on the peak value of excess infection risk to vaccinators. The changes compared to the base case (in Table 1) are shown for the minimum (lighter shading) and maximum (darker shading) parameter values shown in Table 1, and for two different assumptions about PPE effectiveness.

However, in the case of vaccinees and their caregivers, the factors that most influence the excess infection risk differ depending on the effectiveness of PPE (Fig. 4). If PPE is effective at reducing transmission, excess risk is affected most by infections from other community members, which depends on the number of contacts and R_0_. Without effective PPE, excess risk is also influenced by how likely vaccinators are to be infected and how long they remain infectious during the campaign. For example, shorter campaign duration, a shorter infectious period or longer period of latent infection all reduce the chances of vaccinees and caregivers coming into contact with vaccinators infected during the campaign while they are infectious.

**Figure 4.**
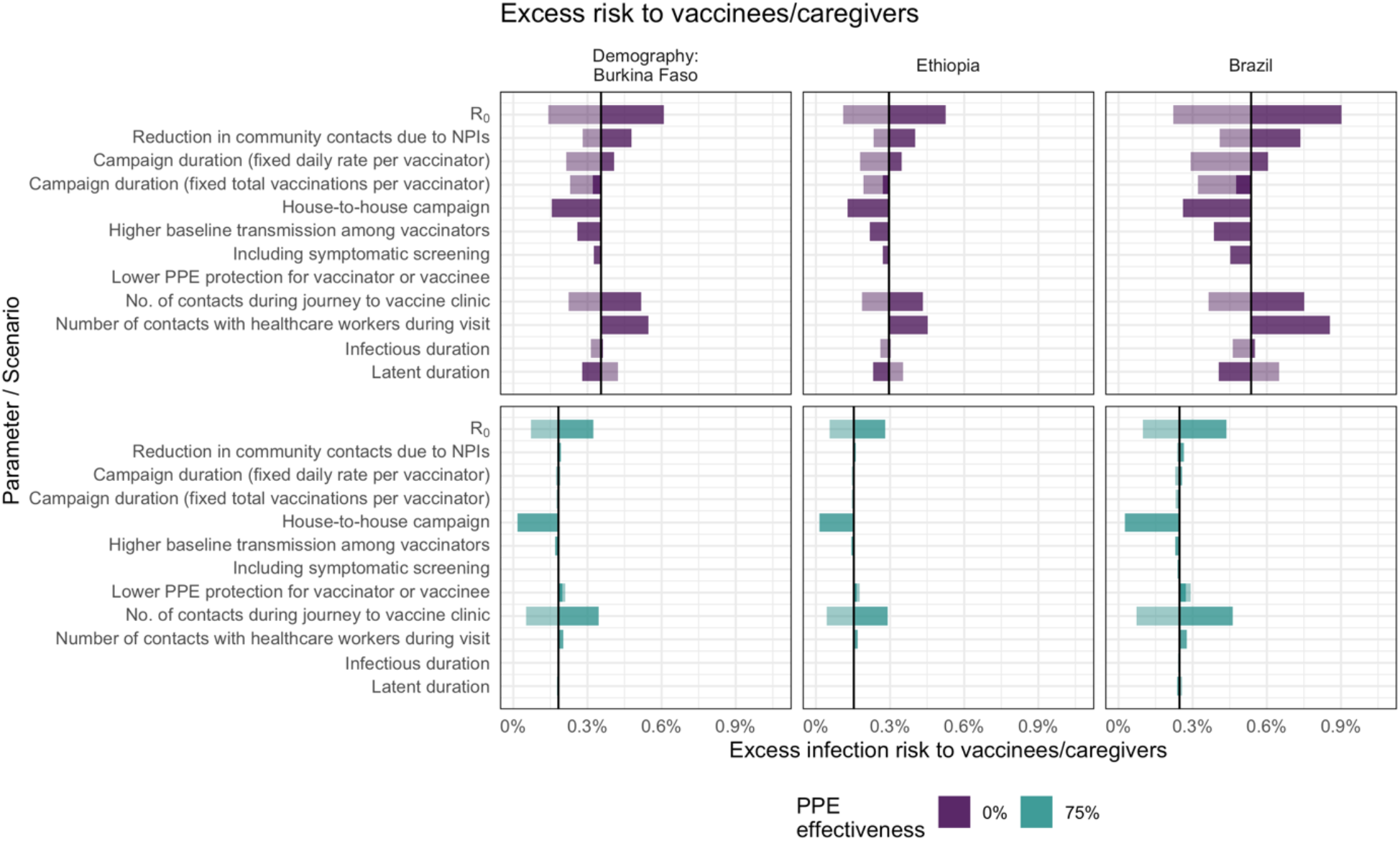
Sensitivity analysis showing the impact of varying model input parameters on the peak value of excess infection risk to children and their caregivers. The changes compared to the base case (in Table 1) are shown for the minimum (lighter shading) and maximum (darker shading) parameter values shown in Table 1, and for two different assumptions about PPE effectiveness.

## Discussion

Using a compartmental transmission model to simulate an epidemic combined with a modified Reed-Frost model, we have estimated the increased infection risk to vaccinators, vaccinated children, and caregivers, of conducting a vaccination campaign during a SARS-CoV-2 outbreak. We find that these risks differ considerably depending on the circumstances in which campaigns are conducted, and the extent to which WHO guidelines are followed.

In the most pessimistic scenarios (e.g. campaigns conducted at the peak of an epidemic without effective PPE), the risks to vaccinators can exceed a 40% increase in infection rate. For vaccinated children and their caregivers, the excess risks are much lower and may be similar to day-to-day infection risk in the general population in lower-risk scenarios (e.g. house-to-house campaigns).

These risks can be dramatically reduced during campaigns, even when community prevalence is high, through the use of effective PPE and by conducting symptomatic screening and isolation of vaccinators. Overall, a campaign at the peak of an outbreak without PPE or screening increases vaccinator risk by an additional 32% to 45% and vaccinee risk by 0.3% to 0.5%. With moderately effective PPE (75%) and screening, this drops to 10% to 15% and 0.15% to 0.25%. At very high levels of effectiveness (90%) consistent with evidence about N95 respirators and other surgical-quality masks, the excess risk for vaccinators can be reduced to 3.6 to 5.3%. These findings support the guidance given by WHO and other associated agencies on the use of adequate PPE and hygiene measures when carrying out campaigns^2^.

For vaccinators, in addition to high community prevalence of SARS-CoV-2, a key factor that increases risk is vaccinating a high number of children during the campaign. However, while the risk for an individual vaccinator can be reduced by vaccinating fewer children, this would require more vaccinators to reach the same size population. We also found that when PPE effectiveness is low a short campaign (and consequently vaccinating many people a day) can reduce infection risk for vaccines and caregivers compared to a longer campaign vaccinating the same number of people overall. However, this assumes that the adequacy of infection control measures is unaffected by campaign duration and the number of individuals vaccinated per day.

Despite the potential for risk mitigation, vaccine campaigns still carry potential for SARS-CoV-2 transmission that needs to be traded off with protecting populations from other vaccine-preventable diseases. These risk-benefit calculations will need to take into account local conditions, and are beyond the scope of this paper, since we have focused only on the risk of COVID-19 itself. However, other studies offer some guiding principles. Routine childhood vaccines should not be interrupted, since the risk of children dying as a result of not receiving these vaccines far outweighs the risk of dying from COVID-19 as a result of attending a vaccine clinic.^10^ For vaccine campaigns, the risk of outbreaks as a result of postponing campaigns varies across settings and pathogens.^11^ For some vaccines (e.g. meningococcal A), campaigns can be postponed in the short-term in many settings due to the persistence of immunity from past campaigns. For other vaccines (e.g. measles), short-term campaign disruptions can lead to large outbreaks very quickly, so campaigns postponed at the height of COVID-19 epidemics should be reinstated as soon as possible afterwards. For polio, countries need to consider the need to avoid jeopardising progress towards eradication as a result of disruption to immunisation activities in 2020.^28^ Hence countries need to consider existing immunity gaps, epidemic potential of diseases and wider initiatives when making difficult decisions about vaccination during COVID-19 outbreaks.

Our study has several limitations in interpretability. First, the actual excess risk to vaccinators depends on their counterfactual risk of infection, which we did not model. If vaccinators are drawn from existing healthcare workers with frequent patient contact, then even if there was no vaccination campaign they would likely be exposed to a high risk of infection without adequate PPE. In fact, if vaccinators have been exposed to a high risk of infection prior to the campaign, this will reduce their excess risk from delivering the campaign, since a large proportion would be either already infected or immune. However, if many vaccinators are already infected, this increases the infection risk for vaccine recipients earlier in the epidemic. Indeed, other analyses have highlighted the role of vaccinators in seeding epidemics in other locations if vaccinators from a high prevalence location deliver vaccination in a lower prevalence location^13^. Furthermore, we did not consider infection risk to staff delivering vaccination campaigns outside of the setting where contact with vaccinees occurs, such as social interactions between vaccination staff or transport of vaccination supplies to clinics.

Second, we did not explicitly assess the contribution of vaccination campaigns to overall infection prevalence in the population. However, even at times of higher prevalence this appears likely to be modest compared to the overall epidemic. This is because only a small portion of the total population are likely to be involved in campaigns (either as vaccinators or vaccine service users.) The impact of vaccination campaigns on excess infection risk for vaccinees and their caregivers is low when compared to the risk to vaccinators and associated delivery staff. However, this is not a rationale to conduct a campaign regardless of the risks if it can safely be postponed, since a similar argument could be applied to any action that has only a marginal impact.

Third, to enhance our study’s generalisability, we modelled hypothetical epidemics rather than actual epidemics experienced in particular settings. In particular, the three exemplar countries (Burkina Faso, Ethiopia, and Brazil) simply represent three examples of population age structure and age-dependent contacts seen in LMICs, rather than actual SARS-CoV-2 epidemiology in the countries. The epidemic of SARS-CoV-2 has varied considerably within these countries, largely due to the impact of non-pharmaceutical interventions, the spread of SARS-CoV-2 variants and increasingly, the impact of COVID-19 vaccination. Modelling the particulars of individual country trajectories was outside the scope of this study. Consequently, assessing the potential impact of vaccination campaigns is best achieved by comparing against the local prevalence of infection in the community.

Fourth, we lacked empirical data on many key drivers of infection risk, such as number of relevant contacts during clinic visits or during travel between households, effectiveness of PPE against transmission, and the baseline transmission risk associated with health-care specific contacts. Many of these parameters were guided either by expert opinion of the vaccine implementers in our advisory group or from single studies in very different settings. Because of this data gap, we varied these parameters across wide ranges in sensitivity analysis, and hence obtained a wide range of potential outcomes. However, if decision makers have access to more specific parameters for their own setting, our open access model could be parameterised to inform their specific decisions.

Fifth, while we used a well-known and extensively validated model of SARS-CoV-2 transmission^14,29,30^, we used a relatively simple extension to capture the risk to vaccinators and vaccine service users during a campaign. In particular, we do not account for the potential campaign-related infections spreading beyond the vaccinators and service users. This was explored by the modelling by Frey et al., who found that most fixed post or house-to-house campaigns do not increase overall infection risks in the general population substantially^13^. This overall conclusion is consistent with our own work, which found that even in the most pessimistic scenarios, overall infection risk to vaccinated children and their caregivers is increased by less than 1.5%. However, Frey et al. highlighted the danger of importing infections to previously naive communities, if vaccinators from one region are moved to another - this effect was not explored in our own work.

Sixth, we assume that asymptomatic or pre-symptomatic vaccinators have the same transmissibility as symptomatic vaccinators, so we could be underestimating the impact of screening if the contribution of asymptomatic transmission (that is not removed by screening) is lower. On the other hand, we assume screening prevents all transmission from symptomatic vaccinators, which likely overestimates screening impact, for example if individuals have atypical symptoms or if adherence to isolation policy is low. Lastly, we did not capture the potential for waning natural immunity, COVID-19 vaccination, or emergence of new variants, which may all become increasingly important in the future.

This analysis raises two important areas of uncertainty in implementation. The first is that risk mitigation of vaccine campaigns is most beneficial when SARS-CoV-2 is low, but accurate estimates of incidence can be challenging in many LMIC settings. While official reporting of country-level cases is valuable, assessment of the local epidemiology may be reliant on anecdotal evidence in some settings. Where possible, more evidence of local incidence is important for detection and response but also to prevent further risks from activities such as vaccine campaigns. The second is that risk mitigation can be very successful with the effective use of PPE, but more specific evidence on what PPE, training and adherence is sufficient to limit risk during vaccine campaigns is lacking.

Despite the limitations of our study, which can be partially addressed by re-parameterisation of our model at a country level, the broad conclusions of our study appear to be robust and are consistent with guidance offered by WHO^5^. In particular, the excess infection risk during a vaccination campaign can be very high, particularly to vaccination staff, but is highly dependent on the timing and characteristics of the campaign, the effectiveness of PPE and the use of symptomatic screening. Other analyses have found that many vaccination campaigns (such as for measles) play an important role in filling susceptibility gaps and preventing outbreaks of vaccine-preventable diseases, and should continue after careful assessment of risks during the COVID-19 pandemic^11^. Hence our results support the use of appropriate risk mitigation measures during campaigns rather than completely calling them off during SARS-CoV-2 outbreaks. Our findings also suggest that staff involved in delivering vaccination campaigns should be considered as potential priority recipients of COVID-19 vaccines alongside other priority groups. Finally, although our investigation specifically focussed on vaccination campaigns, the findings may also be broadly relevant to other campaign-based community interventions, such as vitamin A supplementation and wash activities.

## Conclusion

In conclusion, we find that the SARS-CoV-2 infection risks to vaccinators, vaccinees, and their caregivers from a vaccine campaign conducted during a COVID-19 epidemic can vary considerably depending on the circumstances under which a campaign is conducted. However, these risks can be greatly reduced with effective PPE, symptomatic screening, and appropriate timing of campaigns. Our findings support the continuation of vaccination campaigns using adequate risk mitigation during the COVID-19 pandemic, rather than cancelling them entirely.

## Data Availability

Analysis was performed using R version 4.0.2 and code is available online at: https://github.com/mert0248/vax_campaign_risk

## Declarations

## Acknowledgments

We thank members of our expert advisory group on vaccination programme planning (Svetlana Stefanet, Alaa Rahi, Alexei Ceban, Landry Kaucley and Graça Matsinhe) for helpful discussions about the logistics of vaccination programme delivery in different countries.

The following authors were part of the Centre for Mathematical Modelling of Infectious Disease COVID-19 working group: Naomi R Waterlow, C Julian Villabona-Arenas, James D Munday, Graham F. Medley, Rachel Lowe, Paul Mee, Yang Liu, Amy Gimma, Kevin van Zandvoort, Joel Hellewell, Damien C Tully, Oliver Brady, Megan Auzenbergs, Gwenan M Knight, Adam J Kucharski, Rosanna C Barnard, William Waites, W John Edmunds, Nikos I Bosse, Akira Endo, Emilie Finch, Timothy W Russell, Yung-Wai Desmond Chan, Matthew Quaife, Rosalind M Eggo, Kiesha Prem, Rachael Pung, Thibaut Jombart, Billy J Quilty, Samuel Clifford, Mihaly Koltai, Hamish P Gibbs, Sam Abbott, Christopher I Jarvis, Yalda Jafari, Petra Klepac, Fabienne Krauer, Fiona Yueqian Sun, Sebastian Funk, Frank G Sandmann, Emily S Nightingale, Jiayao Lei, Sophie R Meakin, Alicia Rosello, Carl A B Pearson, David Hodgson, Ciara V McCarthy, Anna M Foss, Katherine E. Atkins.

## Authors’ contributions

SRP, MJ, KA, SF and KMO conceived the study and developed the model. SRP wrote the code, performed the analysis, and generated the figures with guidance from MJ. SRP and MJ wrote the original manuscript. All authors interpreted the results, reviewed and edited the manuscript, and approved the final version.

## Ethics approval and consent to participate

Not applicable.

