## Supplementary Figures for "SARS-CoV-2 infection risk during delivery of childhood vaccination campaigns: a modelling study"

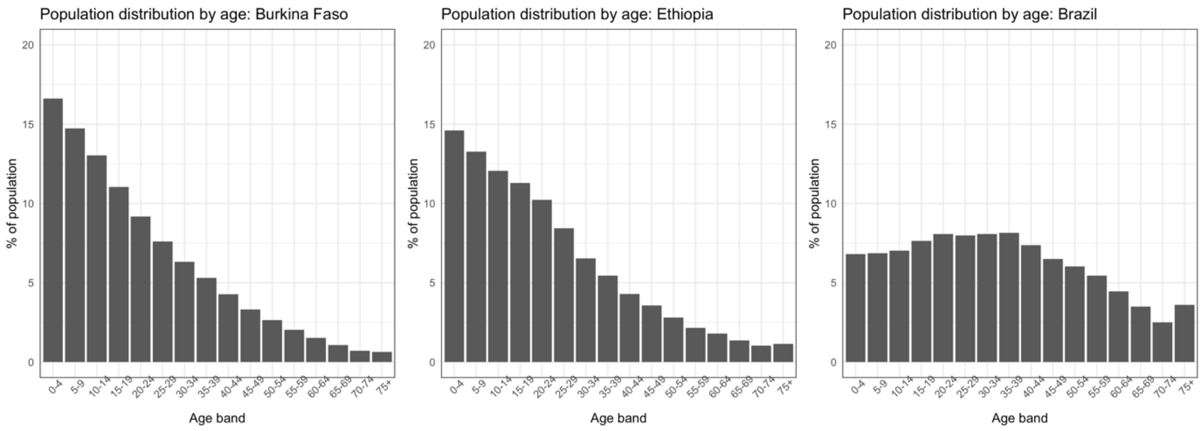


**Supplementary Figure 1. Population age distribution used to parameterise the transmission model**

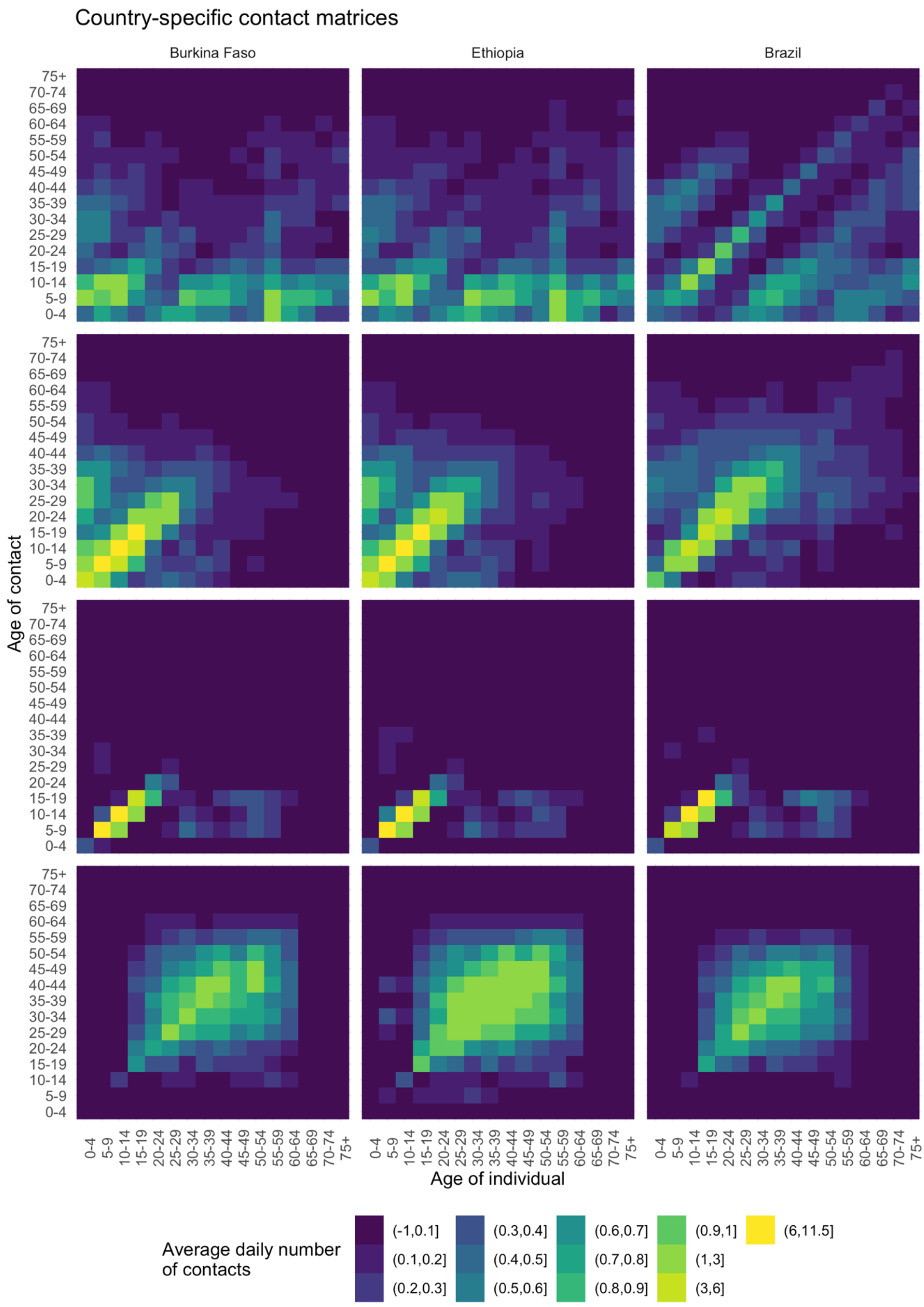


**Supplementary Figure 2. Baseline age-specific contact matrices from Prem et al. (prior to assumed reduction in non-household contacts) used to parameterise the transmission model**

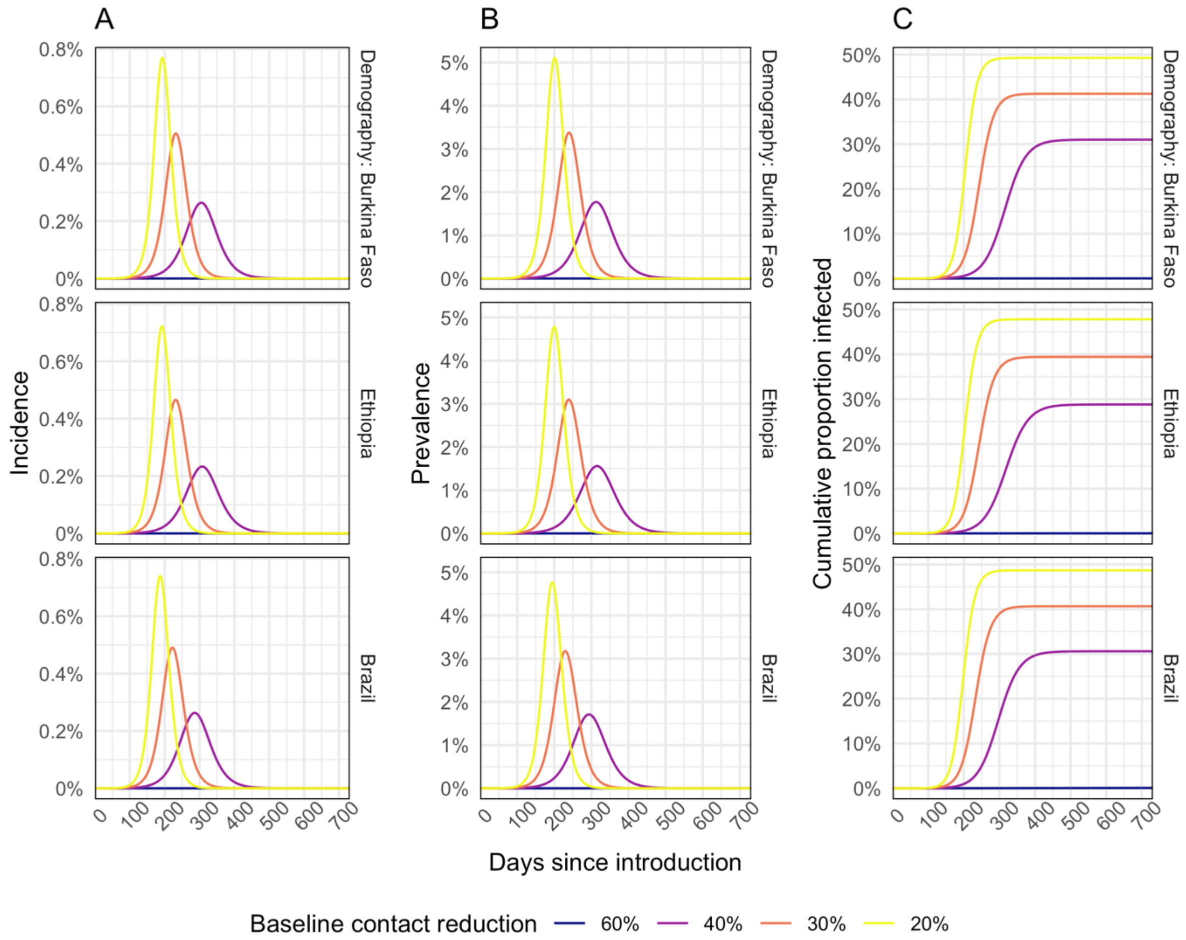


**Supplementary Figure 3. Modelled incidence (A), prevalence (B) and cumulative proportion of individuals infected (C) for an epidemic with $R_0$ = 2 and assuming different % reductions in non-household contacts compared to baseline.** These simulations are used in two different sensitivity analyses: (i) using results for 30% and 20% reductions applied to vaccinators only to model the impact of faster epidemics amongst healthcare workers (Supplementary Figure 4); and (ii) using results for 60% and 20% reductions as a sensitivity analysis applied to the whole population.

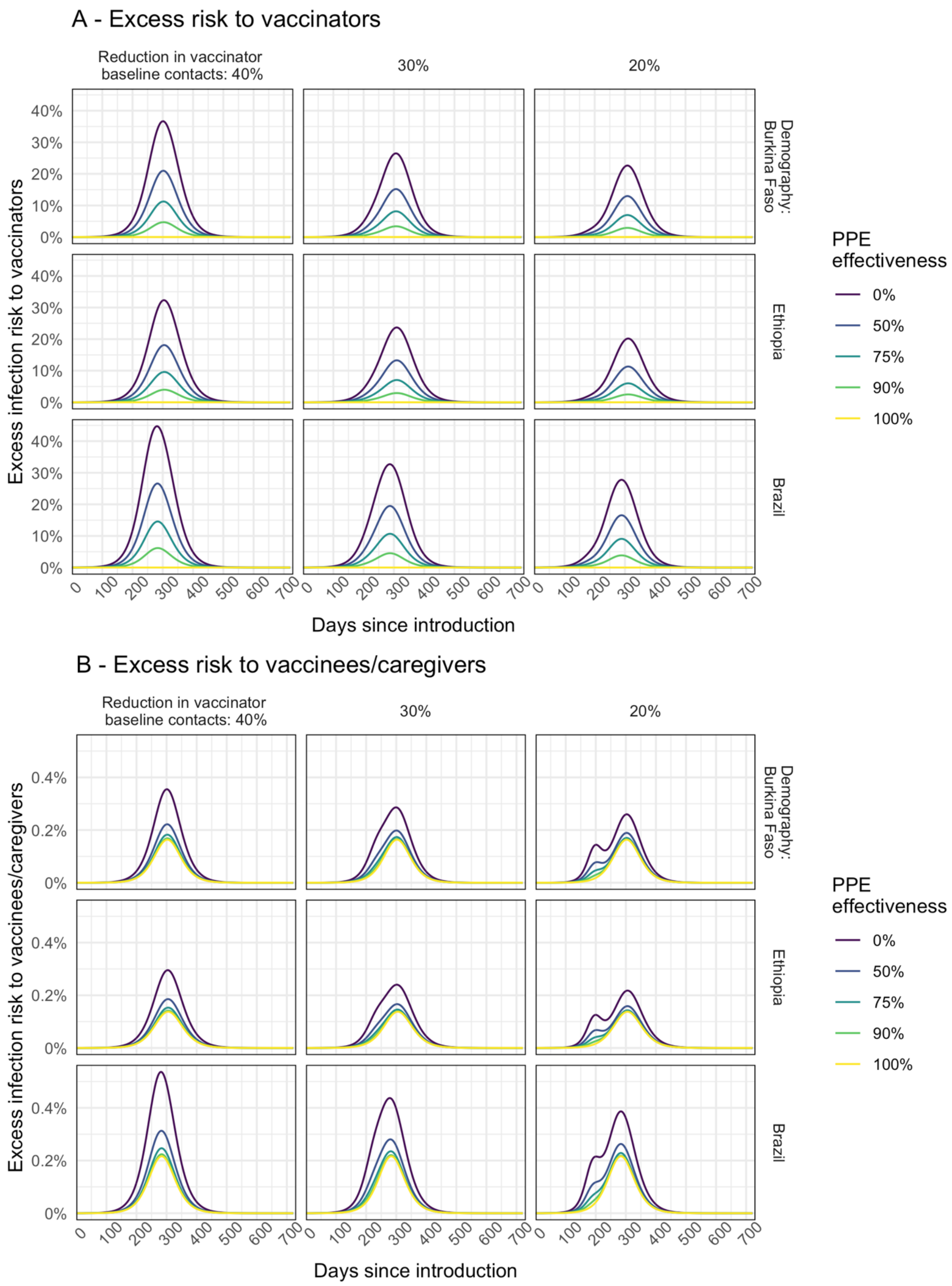


**Supplementary Figure 4. Excess infection risk for vaccinators (A) and children and/or caregivers (B) for scenarios in which vaccinators are assumed to experience a smaller reduction in non-household contacts compared to the general population.**

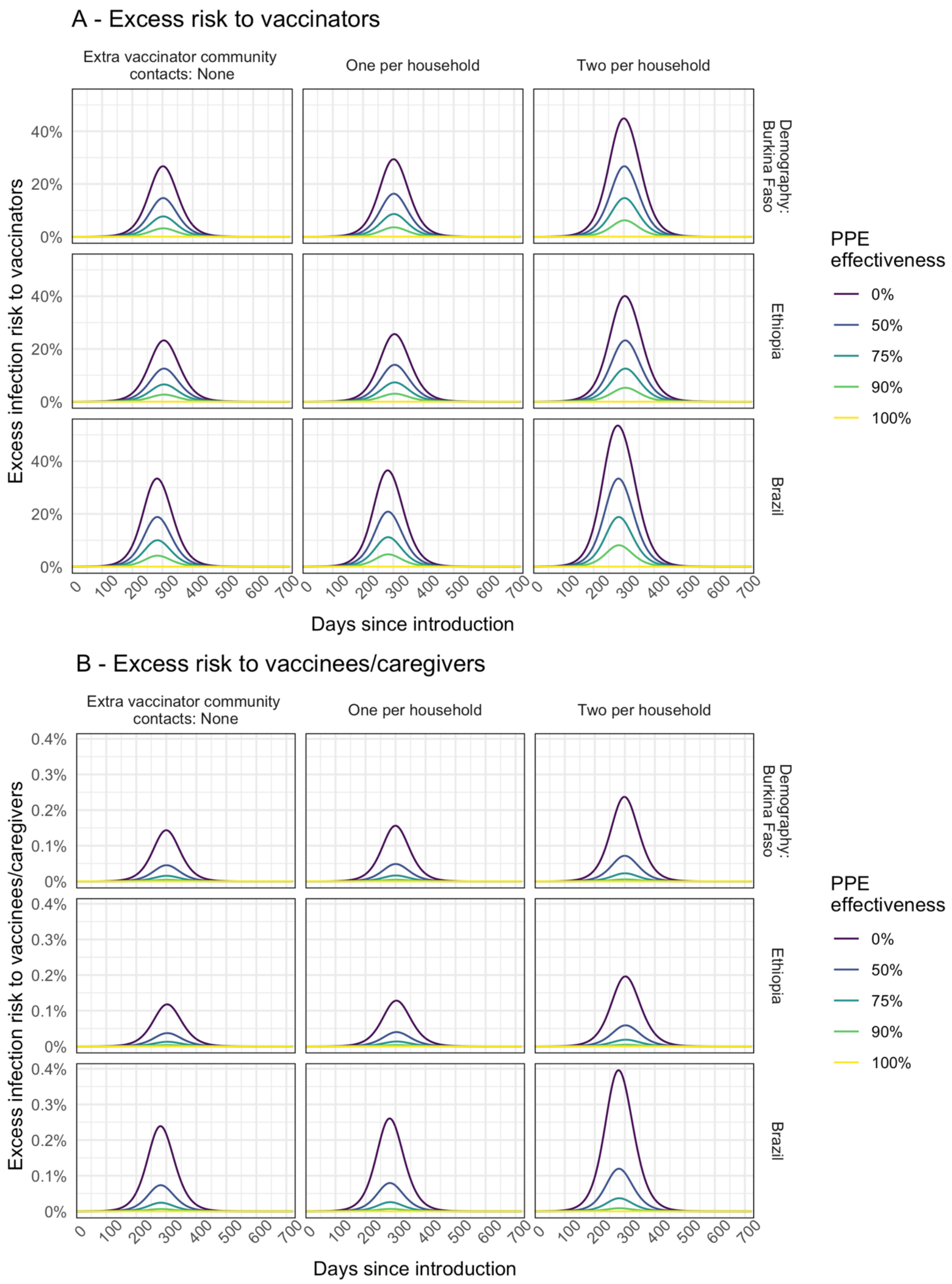


**Supplementary Figure 5. Modelled excess risk of (A) vaccinators and (B) vaccinnees and/or caregivers becoming infected during house-to-house immunisation campaigns conducted at different times during the epidemic.** Results are shown for epidemics modelled using $R_0$=2 and different assumptions about the number of additional community contacts made by vaccinators for each household visited. Line colour shows the impact of different levels of PPE effectiveness.

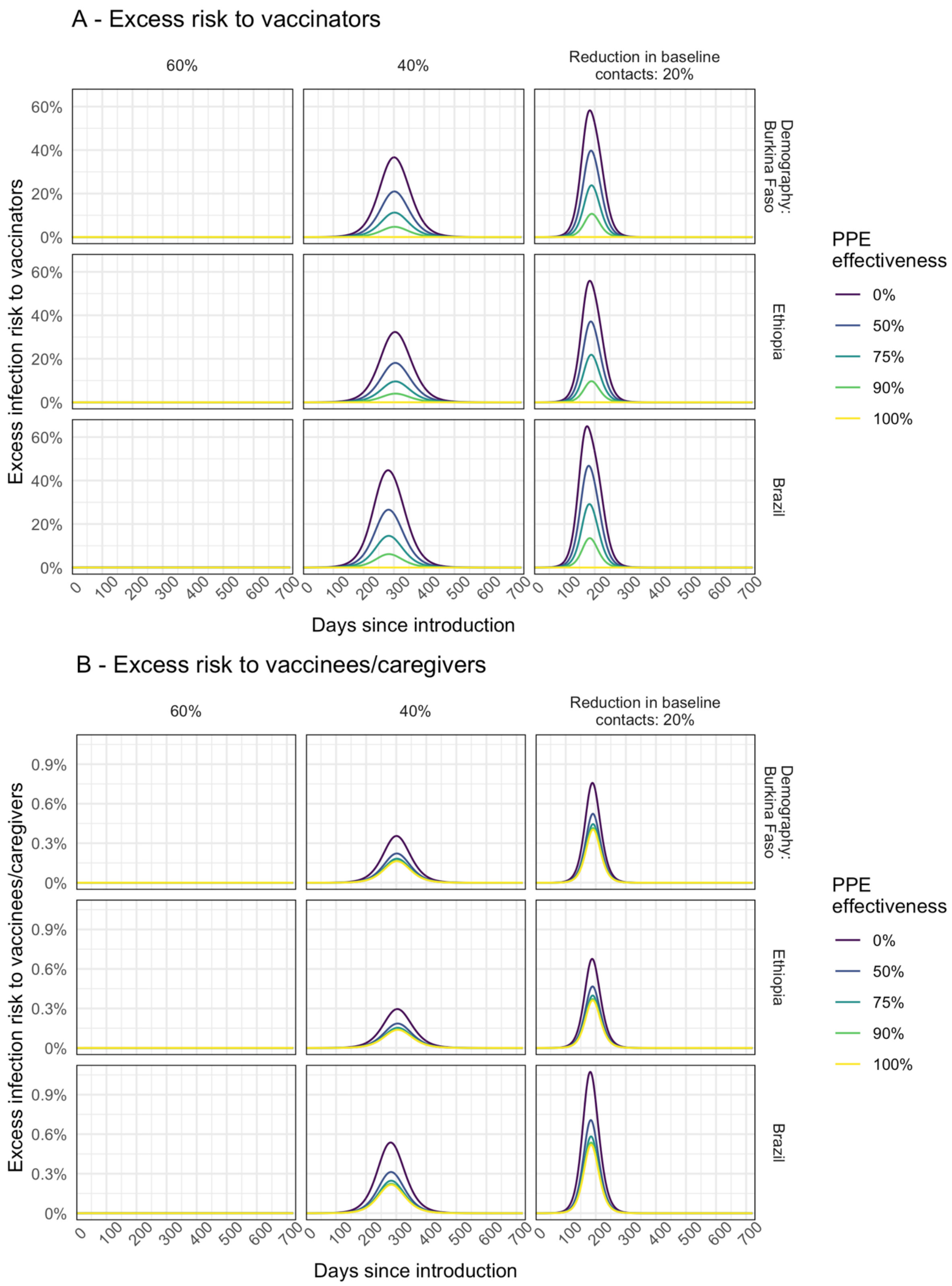


**Supplementary Figure 6. Modelled excess risk of (A) vaccinators and (B) vaccinnees and/or caregivers becoming infected during fixed-post immunisation campaigns conducted at different times during the epidemic.** Results are shown for epidemics modelled using $R_0$=2 and different assumptions about the level of reduction in non-household contacts due to NPIs. Line colour shows the impact of different levels of PPE effectiveness.
